# Negative Symptoms and Their Associations with Other Clinical Variables and Working Memory Across the Schizophrenia Spectrum and Bipolar Disorder

**DOI:** 10.1101/2023.07.18.23292815

**Authors:** Marco De Pieri, Xaver Berg, Foivos Georgiadis, Janis Brakowski, Achim Burrer, Michel Sabe, Mariia Kaliuzhna, Stefan Vetter, Erich Seifritz, Philipp Homan, Stefan Kaiser, Matthias Kirschner

## Abstract

Negative symptoms (NS) of schizophrenia spectrum disorders (SSD) are also prevalent in bipolar-disorder-I (BD-I) and show associations with impaired working memory (WM). However, empirical work on their relationship to other clinical factors across SSD and BD-I are sparse. Here, we characterized the associations of NS with key clinical variables and WM capacity across a combined sample of SSD and BD. We included 50 outpatients with SSD and 49 with BD-I and assessed NS domains using SANS global scores for avolition-apathy, anhedonia-asociality, alogia and blunted affect. We assessed the transdiagnostic relationship between NS and other clinical variables including positive symptoms, disorganization, depressive symptoms, antipsychotic medication using multiple regressions. The strength of these associations was further determined through dominance analyses. Lastly, we used multiple regression to assess the relationship between NS domains and WM. To assess the generalizability of transdiagnostic associations, analyses were repeated in each diagnostic group separately. Across SSD and BD-I, disorganization was associated with avolition-apathy and anhedonia-asociality and depressive symptoms additionally predicted anhedonia-asociality. Antipsychotic dose was associated with blunted affect while group differences only predicted alogia. Higher avolition-apathy was related to impaired WM transdiagnostically, partially mediated by severity of disorganization, whereas only in BD-I higher anhedonia-asociality was associated with better WM capacity. This study demonstrated transdiagnostic associations of both avolition-apathy and anhedonia-asociality with disorganization, and identified avolition-apathy as potential transdiagnostic predictor of WM impairments. Overall, our findings highlight the importance of understanding the relationship between NS domains and other clinical factors with cognitive function across SSD and BD.

## INTRODUCTION

Negative symptoms (NS) are a core feature of schizophrenia spectrum disorders (SSD) and encompass avolition, anhedonia, asociality, alogia and blunted affect^1^. These five symptom domains can be classified in two dimensions: the amotivation dimension includes avolition, anhedonia, asociality and the diminished expression dimension includes blunted affect and alogia^2,3^. Both NS dimensions relate differently to clinical outcomes^4–8^ and have divergent behavioral and neurobiological substrates^3,9–14^. Recent psychometric research supports an even more granular differentiation into the five NS domains^15^ suggesting for example distinct neural correlates of avolition and anhedonia in individuals with SSD. At the neural level, this is supported to date by few studies that identify neural substrates that are distinct for avolition and anhedonia^12^ whereas in most studies neural substrates are associated with both avolition and anhedonia^11,14,16^.

Though NS have been long viewed as a signature symptom of SSD, distinguishing them from other psychotic manifestations such as disorganization and positive symptoms^17^, recent literature suggests NS are also present in other psychiatric conditions^18–20^. Given the clinical, genetic and biological proximity of SSD and bipolar disorder I (BD-I)^21–25^, cross-disorder studies of NS spanning SSD and BD have become increasingly pertinent. Recent studies support such a transdiagnostic construct of NS showing similar levels of avolition and anhedonia but higher levels of blunted affect and alogia in schizophrenia relative to BD-I patients^20,26,27^.

NS can be further distinguished into primary or secondary NS^28,29^. The concept of secondary NS originated in the in the seminal work from Carpenter et al. in the mid-80s^30^, and its significance in contemporary research and clinical practice has been reiterated by various scholars^28,29,31^. Primary NS are thought to be intrinsic to the pathological processes underlying schizophrenia and not related to other symptoms or clinical factors. Secondary NS are defined as NS that are related to other clinical factors including positive symptoms, depression, medications’ side effects, social deprivation, and substance abuse^28–30,32^. In clinical contexts, secondary NS often manifest in various ways: anhedonia may arise from depressive symptoms; avolition can result from pronounced delusions, where delusional beliefs hinder an individual’s capacity to engage in social, occupational, and other goal-directed activities; blunted affect might also be a medication side effect. Such secondary NS might be effectively addressed, if the underlying causes are identified and adequately treated. With respect to NS due to positive symptoms this is indirectly supported by acute treatment trial reporting a joint reduction in positive and NS^33^. In summary, the differentiation of primary and secondary negative symptoms is difficult, in both clinical and research settings. NS that emerge/alleviate in response to changes in causative factors (e.g., onset/recovery of other symptoms; administration/withdrawal of medication) are more likely secondary, while NS that are long-standing and not fluctuating despite changes in causative factors should be considered as primary^29,31^.

Working memory (WM) is the function of temporary storage of information and is critical for multiple other processes including language, executive functions, learning and memory^34^. WM impairments are well documented across all the stages of SSD, from disease onset^35–37^ to the chronic phase^38^. In addition, poorer education and social achievements of patients with SSD have been attributed to WM impairment^39^. Similarly, in patients with BD-I, WM is impaired regardless of the phase of the illness^40^, with deficits persisting in the euthymic phase^41^ and representing the cognitive deficit more specifically related to functional outcomes^42–44^.

Several studies indicate a positive association between cognitive dysfunction, including WM, with negative symptom severity^45–49^ and impaired reward-based decision making^45,50,51^. Moreover, it was proposed that negative symptoms mediate the association between cognitive impairment and functioning in patients with SSD^52^. Recently, attention was given to the transdiagnostic associations between amotivation and impaired WM in both SSD and BD-I^53^.

The present study addressed two intertwined questions to shed light on the cross-disorder significance of different NS domains. The primary objective of the study was to investigate the transdiagnostic relationship between NS domains across SSD and BD and potential underlying sources of secondary NS. The secondary objective of the study was to study the relationship between NS domains and WM function across SSD and BD individuals. Besides these two main objectives focused on a transdiagnostic perspective, we also established an exploratory analysis to test whether the transdiagnostic findings can be confirmed in both SSD and BD separately. Based on previous reports ^26,27^ we hypothesized that levels of avolition and anhedonia would be comparable in SSD and BD-I, while blunted affect and alogia would be stronger in SSD compared to BD-I. For investigation of WM and negative symptoms, we hypothesized that reduced WM capacities would be related to higher levels of avolition/apathy, based on the existing literature ^45–49^.

## METHODS

### Participants

Demographic and clinical data of outpatient individuals with SSD and with BD-I from UCLA consortium for neuropsychiatric phenomics (CNP) were downloaded from the public database OpenfMRI (https://openfmri.org/dataset/ds000030/). In total this database includes a sample size of 50 individuals with SSD and 49 individuals with BD-I, which is line with previous reports investigating negative symptoms across SSD and BD-I^26,27^. The current sample of the CNP was part of a larger multimodal imaging cohort including data from healthy controls and individuals with ADHD, which are not subject to this study. For more details of the CNP cohort see^54^. All the available patients satisfying inclusion and exclusion criteria were recruited, via outreach to local clinics and via online portals. Inclusion criteria were: Age between 21 and 50 years; primary language either English or Spanish; a minimum of 8 years of formal education; no significant medical illness; demonstrated clinical stability allowing to complete the clinical assessment; negative drug test (urine sample tested for cocaine, methamphetamine, morphine, tetrahydrocannabinol, benzodiazepines).

### Clinical assessment

Experienced senior psychiatrists realized a comprehensive clinical assessment was applied across both groups to assess NS and potential sources of NS including positive symptoms, disorganization, depression, and current antipsychotic as well as current mood stabilizing medications. The Scale for the Assessment of Negative Symptoms (SANS) was used to assess the four NS domains blunted affect, alogia, avolition-apathy, anhedonia-asociality. Following previous work from Strauss and Cohen, the global rating scores were used to measure severity of each NS domains^20^. In addition, the motivation and pleasure dimensions were defined by combining the avolition-apathy and anhedonia-asociality domains, and the diminished expression dimension by combining the blunted affect and alogia domains, respectively^5,26^. Positive symptoms were assessed using the Scale for the Assessment of Positive Symptoms (SAPS). Following the work of Peralta et al^55^ and of Tibber et al^17^, global scores of the SAPS domains delusions, hallucinations, bizarre behavior and positive formal thought disorder were used to define two dimensions: the positive symptom dimension includes the delusion and hallucination global scores; the disorganization dimension comprises the bizarre behavior and positive formal thought disorder global scores. To assess depressive symptoms the 21-item version of the Hamilton Scale for Depression (HAMD-21) was used. Lastly, daily antipsychotic doses and use of mood stabilizers were determined. Individual doses of antipsychotics were converted in oral risperidone equivalent doses using the defined daily doses (DDD) method^56^.

### Working memory

WM was assessed using total raw scores from symbol span and visual reproduction (both from the Wechsler Memory Scale), as well as total raw scores from digit span and letter number sequencing (both from Wechsler Adult Intelligence Scale IV). The total raw scores were standardized using z transformation and a composite WM sum score was calculated by combining all four independent z scores. To account for the influence of age on working memory, as previously reported in SSD^57^ and BD-I^58^, age was regressed-out from the composite WM score and the age-corrected residuals were used for all subsequent analysis.

### Data analysis

All analyses were performed using SPSS (version 27.00, SPSS Inc.) and graphics were created with ggplot2 and reshape packages in R (© 2009-2021 RStudio, PBC; 2021.09.0 Build 351).

### Group comparison of clinical variables between SSD and BD-I

Prior to our transdiagnostic analyses, we first applied group comparisons of all clinical variables including positive symptoms, negative symptoms, disorganization, depressive symptoms, daily antipsychotic dose, and use of mood stabilizer. These variables were examined using analysis of covariance (ANCOVA).

### Associations between negative symptoms and other clinical factors

We applied a transdiagnostic approach to examine associations between each NS domain and potential sources of NS across SSD and BD. For each NS domains, we performed a separate multiple linear regression analysis including group, positive symptoms, disorganization, depression, and RSP-eq dose as covariates. Given the body of evidence indicating the possible influence of age^59–64^ and sex^64–68^ on NS, their potential role as confounders was assessed using The Spearman correlation and the Wilcoxon rank sum test for age and sex, respectively. Since these two variables were not significantly related to negative symptoms (**Table S3** and **S4**) in both the transdiagnostic group and in the SSD and BD subgroups, they were not included in the multivariate analysis described below. However, alternative multivariate models including age and sex were also realized to confirm that age and sex did not influence the main findings. We next applied dominance analyses to further contextualize the relative importance of each clinical factor in explaining the NS domains^69^. Dominance analysis is a statistical method designed to establish the relative importance of predictor variables in comparison to all other predictors (in this instance, potential sources of secondary NS) within a statistical model. This method facilitates ranking and scaling predictor variables in multiple regression analyses qualitatively via pairwise comparison. This strategy enables the identification of dominant variables in terms of their efficacy in predicting the variance of the dependent variable in the presence of other variables. **Figure 2** presents a visualization of the relative importance scores for each predictor variable, displayed as a heatmap. Darker shades signify reduced dominance for the respective predictor variable^69^.

To facilitate comparisons with studies employing the two-factor model of NS, all analyses were repeated by executing the multiple linear regression analyses using the two NS dimensions. Finally, to control that none of the transdiagnostic findings was driven by either SSD or BD-I, regression analyses were repeated within each subgroup separately.

### Associations between working memory and negative symptoms

In alignment with the transdiagnostic approach mentioned above, the relationships between WM and NS domains were first assessed across both groups. Considering previous findings of a sex influence on WM in both SSD and BD-I^70–74^, sex was evaluated as a potential confounder in our cohort, but no significant association emerged. Accordingly, sex was not included in the linear regression model for our main results. However, for further precision, a secondary analysis was also realized, including sex in the same regression model. Since age had a significant effect on WM performance in our cohort (as described above), age was first regressed out from the WM composite score. To this end a multiple linear regression model were fitted, using the four NS domains, potential secondary sources of NS, and diagnostic group as independent (predictor) variables. Finally, to assess potential group-specific associations between WM and NS domains, explorative regression analyses were performed in each subgroup separately. We corrected the main transdiagnostic analyses including the four NS domains regression analysis as well as the WM regression analysis (n=5 tests) for multiple comparison using a False Discovery Rate (FDR) of q<0.05 (pFDR <0.05) based on the Benjamini-Hochberg method ^75^.

## RESULTS

### Demographic and clinical data

Fifty patients with SSD and 49 patients with BD-I were included in the study; All participants were outpatients and diagnoses were confirmed following DSM-IV criteria using a semi-structured assessment with Structured Clinical Interview (SCID) for the DSM-IV (**Table S1**). Demographic and clinical data are summarized in in **Table 1**. We observed no group difference in age (F = 0.128, p = 0.880), but the proportion of male subjects in the SSD group compared to the BD-I group was higher (SSD (76%) vs BD-I (57%), *χ*^2^=11.00; p < 0.001). Both groups exhibited mild to moderate levels of SANS avolition-apathy and SANS anhedonia-asociality and subclinical levels of SANS blunted affect and SANS alogia, with very low levels of alogia in BD-I.

**Table 1.** Demographic and clinical data.

|  | SSD (N=50) | BD-I (N=49) | Test-statistic |
| --- | --- | --- | --- |
| Age (years) | 36.46 ± 8.878 | 35.61 ± 9.140 | F = .128 (p = .880) |
| Sex (male / female) | 38 / 12 | 28 / 21 | $\chi^2 = 3,960$ (p = 0.047) |
| Ethnic group |  |  |  |
| American Indian or Alaskan Native | 11 (22%) | 4 (8.2%) |  |
| Asian | 1 (2%) | / |  |
| Black/African American | 2 (4%) | 1 (2%) |  |
| White | 33 (66%) | 37 (75.5%) |  |
| More than one race | 1 (2%) | 7 (14.3%) |  |
| SANS |  |  |  |
| Apathy-Avolition | 2.72 ± 1.512 | 1.85 ± 1.382 |  |
| Anhedonia-Asociality | 2.28 ± 1.499 | 1.70 ± 1.245 |  |
| Alogia | .98 ± 1.152 | .15 ± .420 |  |
| Blunted affect | 1.26 ± 1.306 | .57 ± 1.088 |  |
| Amotivation | 5.00 ± 2.770 | 3.54 ± 2.287 |  |
| Diminished expression | 2.24 ± 2.246 | .72 ± 1.393 |  |
| SAPS |  |  |  |
| Positive symptoms | 4.84 ± 2.795 | .72 ± 0.98 | F = 44.443 (p = .000) |
| Disorganization | 2.56 ± 2.28 | 1.63 ± 1.77 | F = 2.911 (p = .059) |
| HAMD-21 | 11.92 ± 9.001 | 13.72 ± 9.75 | F = 3.765 (p = .027) |
| Risperidone equivalent dose (mg/d) | 7.48 ± 8.79 | 4.85 ± 10.93 | F = 1.41 (p = 0.25) |
| Patients with mood stabilizer intake | 11 (22%) | 30 (61%) | $\chi^2 = 15,69$ (p < 0.001) |

*SSD, schizophrenia spectrum disorder; BD-I, bipolar disorder I; Test-statistic, ANCOVA (sex as covariate); SANS, Scale for the assessment of negative symptoms. SANS amotivation includes the SANS domains avolition-apathy and anhedonia/asociality. SANS diminished expression includes the SANS domains alogia and blunted affect. SAPS, Scale for the assessment of positive symptoms. SAPS positive symptoms includes the SAPS domains delusion and hallucination. SAPS disorganization includes the SAPS domains bizarre behavior and positive formal thought disorder; HAMD-21, Hamilton Depression Scale (21 items)*

With respect to other symptom dimensions, subjects with SSD had higher SAPS positive symptoms dimension scores (F = 44.443, p = 0.000) than subjects with BD-I, while subjects with BD-I had higher HAMD-21 scores (F = 3.765, p = 0.027). In contrast, SAPS disorganization dimension scores did not differ between groups (F = 2.911, p = 0.059). Comparing medication dose, we found no group differences in risperidone equivalent doses (F = 1.41, p = 0.25), while mood stabilizers were more often prescribed in the BD-I group (*X*^2^ = 15,69, p < 0.001).

### Potential clinical sources of negative symptoms

Multiple linear regression models were used to contextualize potential sources for negative symptoms for each NS domain across SSD and BD-I. Both higher SANS avolition-apathy (β = 0.25, p < 0.001), and higher SANS anhedonia-asociality (β = 0.20, p < 0.005) were significantly associated with SAPS disorganization dimension, while depressive symptoms were additionally related to SANS anhedonia-asociality (β = 0.05, p < 0.001). Risperidone equivalent dose was associated with higher SANS blunted affect (β = 0.03; p < 0.05), but this result was not significant after correction for multiple testing. SANS alogia showed no relationship with any of the potential sources for secondary NS. Positive symptoms were assessed as potential secondary sources, but no significant associations between the SAPS positive dimension and any NS domain were observed across both groups (**Figure 1**, **Table 2**). Notably, significant group differences were only observed in alogia (β = −0.74; p < 0.05) with more severe alogia in SSD compared to BD-I. To further quantify the variance of NS explained through each clinical factor related to NS, we performed separate dominance analyses for each NS domain. The generalized relative importance values are shown in **Figure 2**, indicating how the underlying sources of secondary NS contribute to the measured SANS domain scores. Overall, the dominance analyses confirm the findings of the multiple linear regression models, highlighting the importance of disorganization in predicting SANS avolition-apathy and anhedonia-asociality.

**Figure 1.**
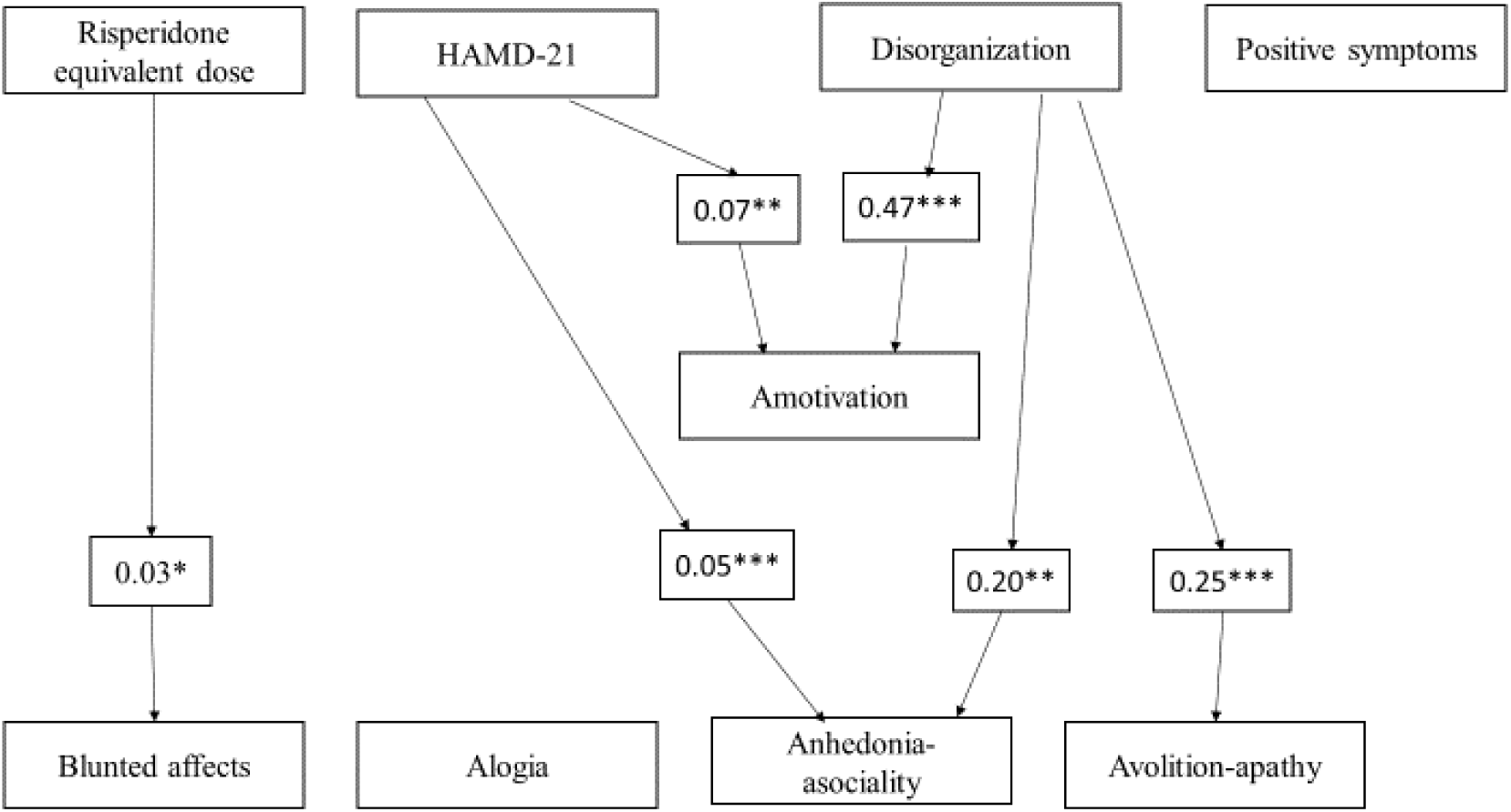
Associations between Negative symptoms and their potential sources. Significant beta values from linear multivariate regressions shown in Table 2 are presented. *p<0.05; **p<0.005; ***p<0.001

**Figure 2.**
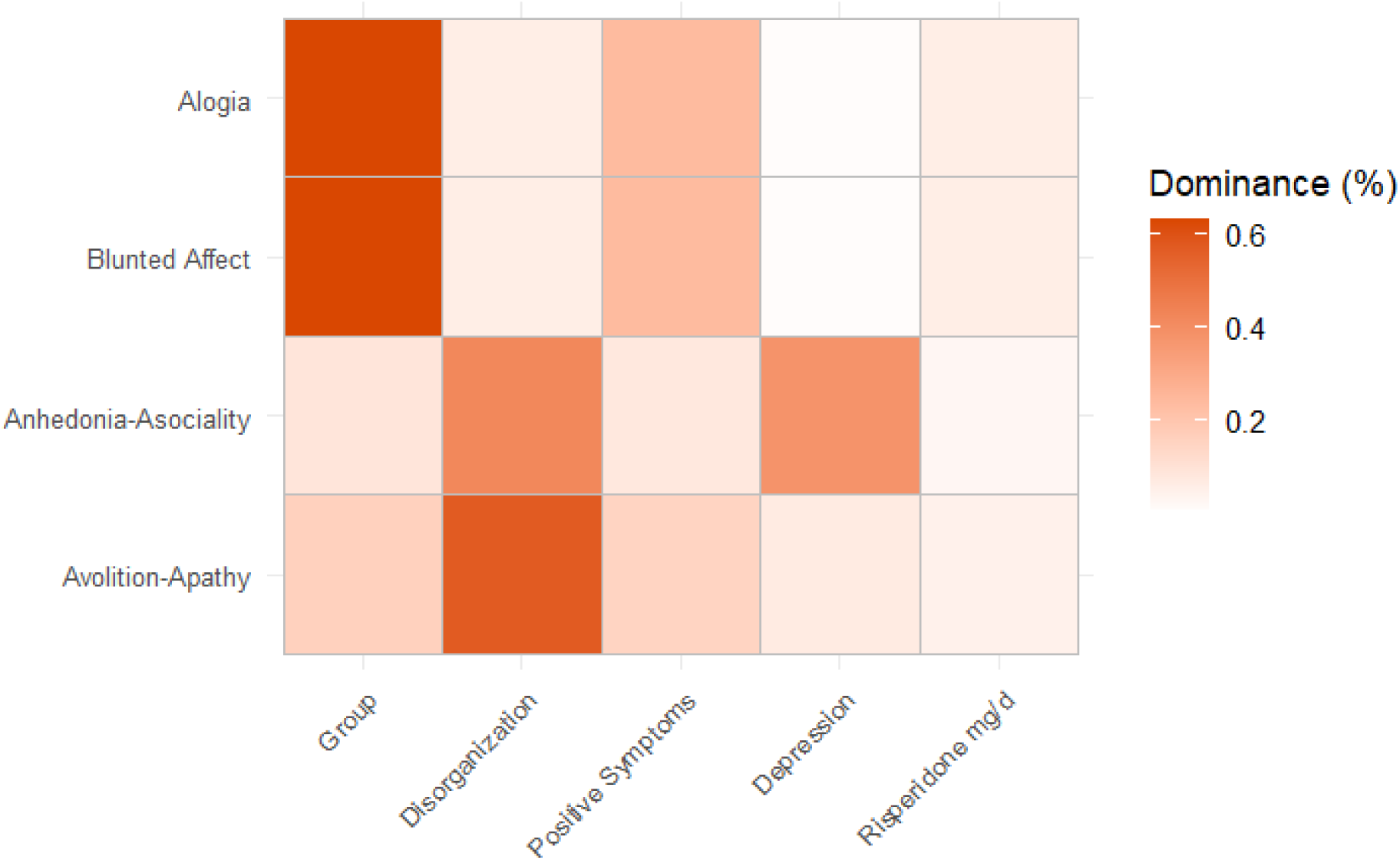
Dominance analysis of potential secondary sources of the negative symptom domains. Visualization of relative importance scores of causes of secondary negative symptoms on a standardized scale from 0 (no importance) to 1 (high importance) as a heatmap. Brighter colors are corresponding to higher percentage of relative importance.

**Table 2.** Relationship between negative symptoms domains and other clinical factors.

|  | **Beta [95% CI]** |  |  |  |
| --- | --- | --- | --- | --- |
|  | Avolition-Apathy | Anhedonia-Asociality | Alogia | Blunted affect |
| **Model fit** | $r^2_{\text{adj}} = .19$<br>$F(5,91) = 5.53^{***}$ | $r^2_{\text{adj}} = .26$<br>$F(5,90) = 7.66^{***}$ | $r^2_{\text{adj}} = .14$<br>$F(5,91) = 4.26^*$ | $r^2_{\text{adj}} = .13$<br>$F(5,91) = 3.80^*$ |
| **Group** | -0.52 [-1.37 – 0.29] | -0.58 [-1.32 – 0.16] | **-0.74[-1.28 - -0.19]\*\*** | -0.53 [-1.24 – 0.18] |
| **Positive Symptoms** | 0.007 [-0.14 – 0.16] | -0.03 [-0.16 – 0.10] | 0.004[-0.09 – 0.10] | 0.03 [-0.01 – 0.16] |
| **Disorganization** | **0.25[0.11 – 0.40]\*\*\*** | **0.20 [0.07 – 0.34]\*\*** | 0.04[-0.06 – 0.13] | 0.008 [-0.12 – 0.13] |
| **Depression** | 0.02 [-0.012 – 0.05] | **0.05 [0.03 – 0.08]\*\*\*** | 0.001[-0.02 – 0.02] | 0.02[-0.003 – 0.05] |
| **RSP-eq dose** | 0.02 [-0.01 – 0.05] | 0.01 [-0.01 – 0.04] | 0.008[-0.01 – 0.03] | 0.03[0.004 – 0.05]* |

*Linear multivariate regressions of 4 negative symptom domains as dependent variables with potential sources of negative symptoms as independent variables across SSD and BD-I patients. Beta values and corresponding 95% confidence intervals are given; Positive symptoms were measured using the Scale for the assessment of positive symptoms (SAPS); Depressive symptoms were measured using the Hamilton Depression Scale (21 items); RSP-eq dose, daily antipsychotic dose as risperidone equivalent; \* $p < 0.05$ \*\* $p < 0.005$ \*\*\* $p < 0.001$ . Statistically significant results after correction for multiple testing (Benjamini-Hochberger procedure, false discovery rate 5%) are bolded.*

Using the SANS, amotivation and the diminished expression dimensions largely replicated the results from the four SANS domain scores. SAPS disorganization scores and depressive symptoms were associated with the SANS amotivation dimension (β = 0.47, p < 0.001 and β = 0.07; p < 0.05, respectively). In line with the observed group differences in SANS alogia, the SANS diminished expression was different in SSD and BD-I (β = −1.27, p < 0.05; SSD>BD-I) (**Table 3**). Finally, analyses for SSD and BD-I separately, largely confirmed the cross-disorder results, albeit with smaller effect sizes (**Table S5**). Taken together across SSD and BD-I, disorganization, depressive symptoms and antipsychotic medication were significantly associated with NS domains, while group differences only predicted alogia. Finally, repeating the analyses including age and sex in the regression model, did not significantly change the results (**Supplementary Table S6**).

**Table 3:**
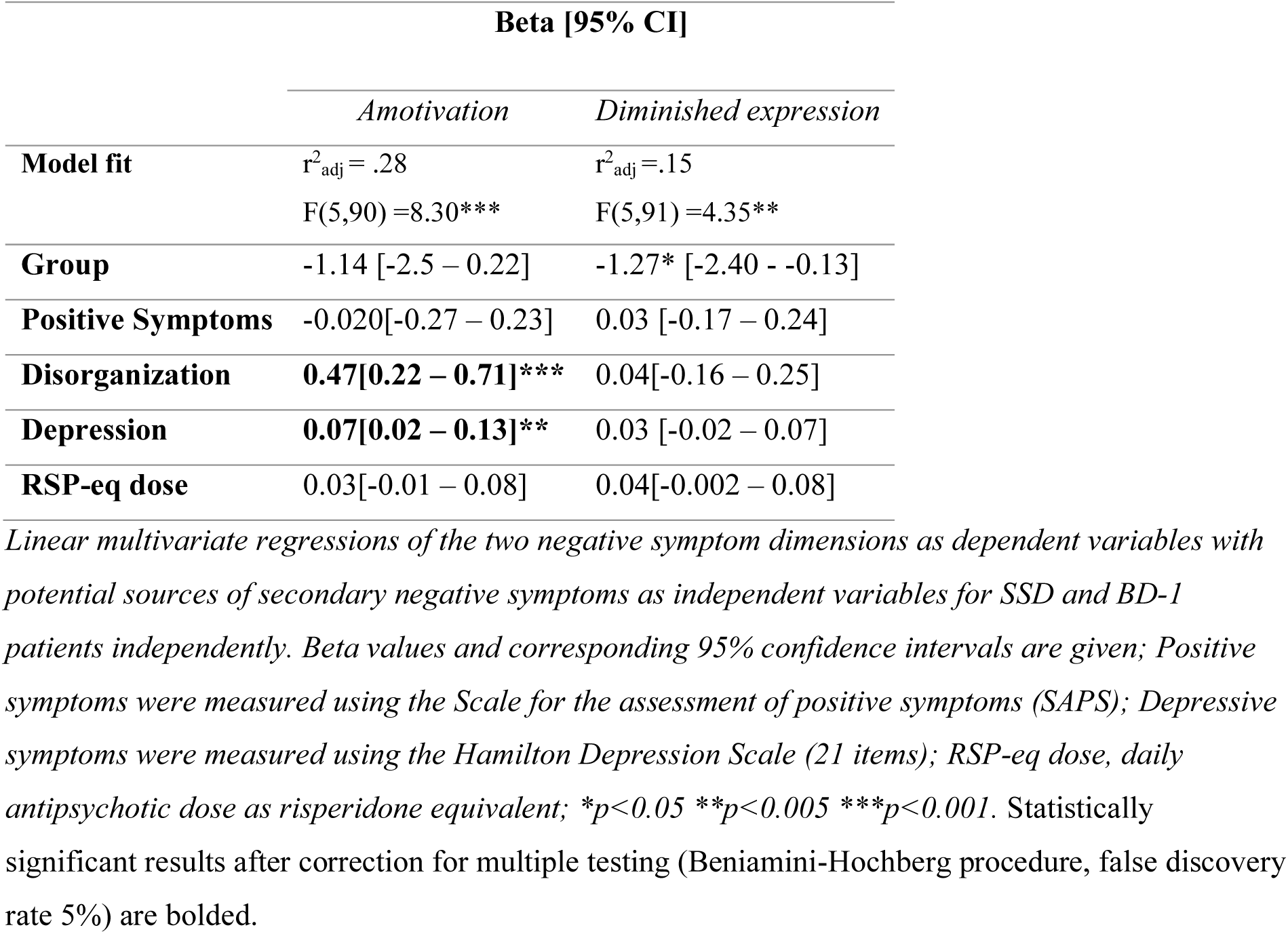
Model fit and negative symptom dimensions scores and potential sources of secondary negative symptoms all over schizophrenia spectrum and bipolar disorder.

### Associations of working memory and clinical variables

In a next step, we sought to examine the relationship between NS and WM across SSD and BD-I and in each group separately (**Table 4**). In our transdiagnostic model, higher WM scores were explained by group (β = 2.77, p < 0.005) with better WM in BD-I than SSD. Furthermore, across both groups, a higher SANS avolition-apathy scores were associated with lower WM scores (β = −0.74, p < 0.005). None of the other symptom dimensions or medication dose were related to WM. Examining the relationship between NS and WM in SSD and BD-I separately, the negative effect of SANS avolition-apathy was observed in both disorders although with higher effect size in individuals with BD-I (β = −1.07, p < 0.001) compared to individuals with SSD (β = −0.58, p > 0.05). In addition to the negative relationship between higher SANS avolition-apathy scores and WM, subjects with BD-I showed an inverse positive association between higher SANS anhedonia-asociality and WM (β = 1.32, p < 0.005). In other words, BD-I subjects with higher SANS avolition-apathy had reduced WM scores, while those with higher SANS anhedonia-asociality had higher WM scores. In sum, across both groups more severe SANS avolition-apathy scores and group differences (SSD<BD-I) predicted lower WM, while in BD-I only, higher SANS anhedonia-asociality showed an inverse relationship with WM (i.e. higher scores). **Supplementary table S7** shows the results obtained with a different regression model including sex among the predictive variables, which led to no relevant differences compared to the results displayed in this paragraph.

**Table 4.** Relationship between clinical variables and working memory.

|  | **Transdiagnostic** | **SSD** | **BD-I** |
| --- | --- | --- | --- |
| **Model fit** | $r^2_{\text{adj}} = .27$<br>$F(9,86) = 4.93^{***}$ | $r^2_{\text{adj}} = -.02$<br>$F(8,41) = 0.85$ | $r^2_{\text{adj}} = .27$<br>$F(8,37) = 3.09^{**}$ |
| **Group** | 2.77[1.14 – 4.40]** | / | / |
| **Avolition-Apathy** | **-0.74[-1.22 - -0.27]\*\*** | -0.58[-1.38 – 0.23] | -1.07[-1.63 – -0.50]*** |
| **Anhedonia-Asociality** | 0.46[-0.1 – 1.03] | 0.01[-0.82 – 0.85] | 1.32[0.40 – 2.24]** |
| **Alogia** | -0.40[-1.19 – 0.39] | -0.47[-1.47 – 0.54] | 1.10[-1.9 – 4.08] |
| **Blunted affect** | -0.05[-0.65 – 0.55] | 0.03[-0.87 – 0.92] | -0.55[-1.68 – 0.58] |
| **Positive symptoms** | 0.17[-0.11 – 0.46] | 0.12[-0.23 – 0.46] | 0.61[-0.26 – 1.47] |
| **Disorganization** | -0.018 [-0.32 – 0.28] | 0.11[-0.33 – 0.55] | -0.12[-0.59 – 0.35] |
| **Depression** | -0.007[-0.07 – 0.06] | 0.03[-0.08 – 0.14] | -0.10[-0.20 – 0.002] |
| **RSP-eq dose** | -0.023[-0.08 – 0.03] | 0.03[-0.07 – 0.126] | -0.04[-0.12 – 0.03] |

*Linear regressions of negative symptom domains and other clinical variables as independent variables and working memory sum score as dependent variable across SSD and BD-I patients. Beta values and corresponding 95% confidence intervals are given; Positive symptoms were measured using Scale for the assessment of positive symptoms (SAPS); depressive symptoms were measured using Hamilton Depression Scale (21 items); RSP-eq dose, daily antipsychotic dose as risperidone equivalent; \* $p < 0.05$ \*\* $p < 0.005$ \*\*\* $p < 0.001$ . Statistically significant results after correction for multiple testing (Benjamini-Hochberg procedure, false discovery rate 5%) are bolded. Correction for multiple testing was not applied to SSD and BD-I subgroups.*

### Mediation analysis

Poor WM is associated with both disorganization^76,77^ and avolition-apathy^4,19^ in patients with SSD. In the given sample, higher disorganization correlated with higher avolition-apathy and higher avolition-apathy corresponded to lower WM. Therefore, we performed a mediation analysis to examine whether disorganization influences the effect of WM on avolition-apathy (see Figure 3). Regressing disorganization on WM showed a trend effect of higher WM being associated with lower disorganization scores (a). Regressing disorganization on avolition-apathy revealed a highly significant positive association (b). When regressing WM on avolition-apathy (c), a negative effect was observed, with lower WM correlating with higher avolition-apathy. Adding disorganization as a mediator (c’) reduced the significant association between WM and avolition-apathy (c) and showed a significant mediation effect of disorganization on the relationship between WM and avolition-apathy (a*b). Collectively, this analysis suggests that WM has a significant direct effect on avolition-apathy and an additional indirect effect mediated by disorganization across SSD and BD.

**Figure 3.**
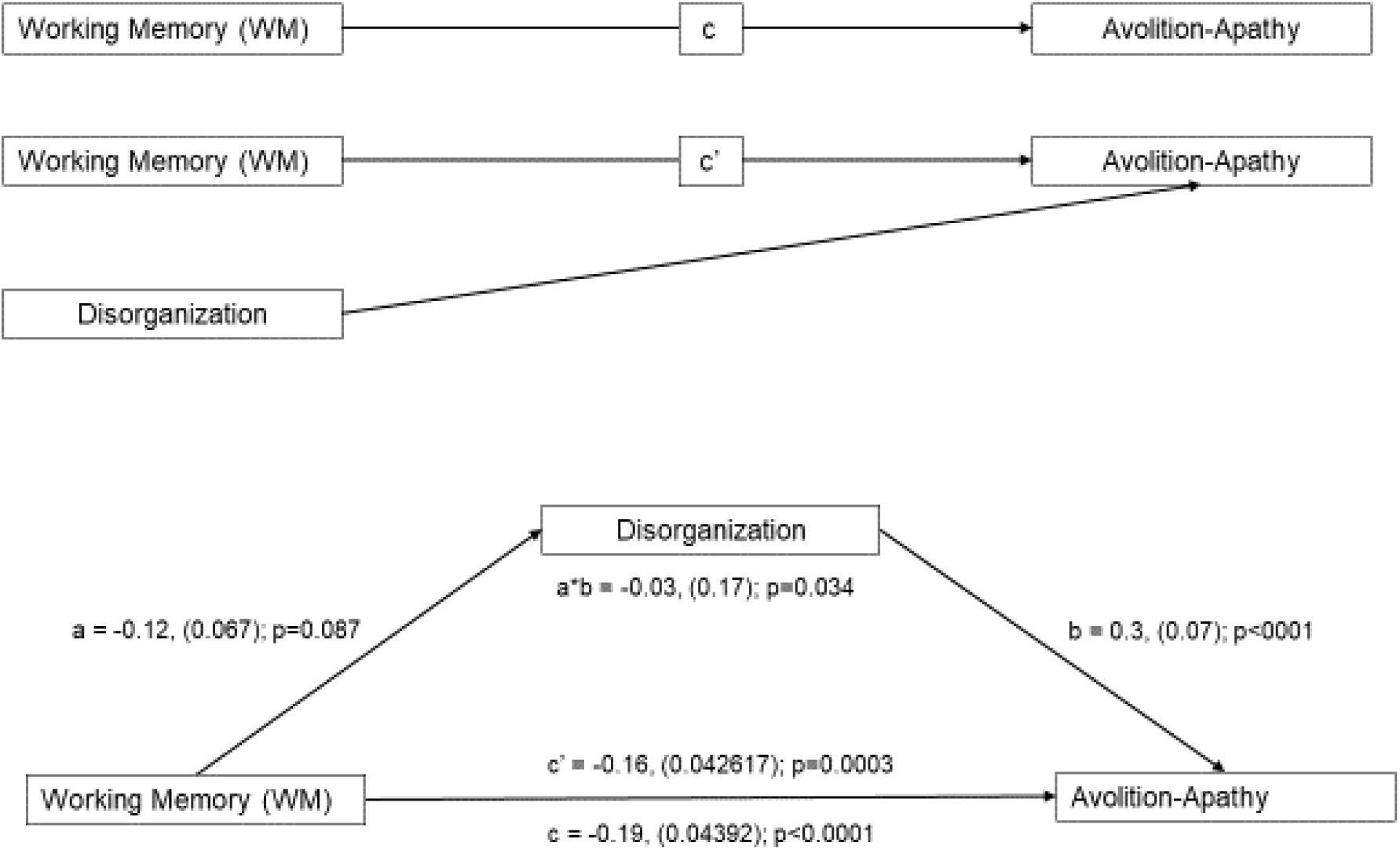
Mediation analysis of working memory (WM), disorganization and avolition-apathy.

## DISCUSSION

In the present study we applied a transdiagnostic approach to characterize NS domains and their relationships with potential sources of secondary NS across a combined sample of SSD and BD-I. We further evaluated the transdiagnostic association between distinct NS domains and WM capacity. Across individuals with SSD and BD-I, disorganization was associated with avolition-apathy and anhedonia-asociality; depression additionally predicted anhedonia-asociality. In addition, a higher antipsychotic dose was associated with blunted affect. Among all four NS domains only alogia was better predicted by disease categories and not by transdiagnostic clinical dimensions. Avolition-apathy was associated with a reduced WM performance transdiagnostically, while only in the BD-I group higher anhedonia-asociality was associated with better WM. Collectively, these findings provide evidence for associations between NS and disorganization and reveal differential effects of avolition-apathy and anhedonia-asociality on WM in SSD and BD-I.

Over the last years NS have been increasingly recognized as a transdiagnostic construct^20^, although if, to our knowledge, only few studies have directly compared NS between SSD and BD-I. With respect to the different NS domains, similar levels of avolition and anhedonia have been reported for SSD and BD-I^26,27^. In contrast, alogia^26,27^ and blunted affect^27^ differed between both groups with significantly more severe scores in SSD compared to BD-I. Together with these reports, the present findings suggest that avolition and anhedonia are transdiagnostic domains across SSD and BD-I. The diminished expression dimension and in particular the severity of alogia, however, seems to be more strongly differentiated by disease category. Secondary NS are frequently observed in clinical routine; however, empirical studies examining potential underlying sources of secondary NS remain relatively scarce. In the present study higher disorganization was associated with higher avolition-apathy as well as anhedonia-asociality across both disorders and confirmed in each disorder separately. Disorganization has been conceptualized as one core dimension^78^ along with positive symptoms and NS, which has been consistently confirmed in SSD^17,55^ and across the SSD-BD spectrum^25^. However, the relation between disorganization and NS has not been receiving much attention. The present findings suggest that disorganization might be a source of secondary avolition-apathy/anhedonia-asociality in patients of the SSD-BD spectrum. One clinical explanation for this association might be that disorganization directly affects the patient’s ability to plan and engage in activities and social interaction. In addition, the relationship between disorganization and NS could be of direct clinical interest. In fact, preliminary evidences exist that cariprazine, a 3^rd^ generation antipsychotic drug, could improve NS and additionally disorganization in SSD patients^79^. Hence, secondary avolition-apathy and anhedonia-asociality due to disorganization might partially respond to a treatment with a 3^rd^ generation antipsychotic like cariprazine. It is important to note here that it is also possible that there is no causal relationship between disorganization and NS. Thus, the potential shared treatment effect of cariprazine may be the result of a common underlying mechanism that is targeted by this drug and improves both symptoms independently.

Anhedonia-asociality was additionally related to depressive symptoms across both groups, which is in line with anhedonia as shared feature of depressive symptoms and NS in SSD and BD-I^26,80,81^. With respect to medication effects, higher daily antipsychotic doses were associated with blunted affect across both disorders. The association, however, was mainly driven by the BD-I group, although medication dose did not differ between BD-I and SSD patients. One previous study reported a similar relationship in BD-I patients but not SSD patients^27^, raising the question whether BD-I patients are more sensitive to medication-induced affective flattening compared to SSD patients. Extrapyramidal side effects are often suggested as cause of diminished expressivity^82^, although other mechanism might contribute to this relationship. Collectively, these findings show that antipsychotic medication relates to even mild to moderate blunted affect that could potentially be ameliorated with precise drug monitoring. Of all four NS domains tested in this study, only alogia was predicted by diagnostic category showing significant higher scores in SSD compared to BD-I patients. In addition, alogia was not related to any potential secondary sources, thus suggesting that alogia was a primary NS in this cross-disorder sample. This observation raises the question whether some NS domains are more likely to be secondary due to other clinical factors, while others such as alogia are more likely primary NS. Neurocognitive deficits are common in psychotic disorders^83^ and in particular WM capacity is typically impaired in both, patients with SSD or BD-I^84^. Impaired WM has been further related to NS ^52^as well as disorganization^76,77^ in patients with SSD, but not in BD-I patients.

We observed that across SSD and BD-I patients, lower WM was associated with higher avolition-apathy scores suggesting a direct effect of WM on avolition-apathy. Disorganization partly mediated the effect of WM on avolition-apathy, pointing to an additional indirect effect of WM on avolition-apathy. The negative effect of primary avolition-apathy on WM has been consistently observed in different stages of SSD^4,19,53,85^. Our findings extend these reports by showing that avolition-apathy, associated with disorganization, relates to more severe WM deficits across SSD and BD-I patients. Of note, the relationship between higher avolition-apathy and lower WM performance was more pronounced in BD-I confirming that this result is not purely driven by the SSD group. These results further provide evidence for a transdiagnostic link between WM and higher avolition-apathy scores, which is shared across the SSD-BD spectrum.

In contrast to this transdiagnostic relationship, we observed an inverse association of higher anhedonia-asociality with better WM in BD-I patients only. Anhedonia-asociality was also associated with depressive symptoms in both diagnostic groups. Therefore, one interpretation is that those BD-I patients with subclinical affective symptoms showed overall better WM performance compared to those with less affective symptoms. In addition, this observation was specific for BD-I patients and could potentially reflect the overall better neurocognition seen in BD-I compared to SSD. This interpretation would be in line with a general model of cognitive functioning across the schizophrenia-bipolar spectrum showing that patients with more affective symptoms have overall better cognitive functioning compared to patients with primary and more enduring psychotic symptoms^86^.

### Limitations

There are several limitations to our study. First, the SANS was used to assess NS. SANS is a first-generation questionnaire for the assessment of NS and newer scales such as the Brief Negative Symptom Scale or the Clinical Assessment Interview for Negative Symptoms are available. However, according to the EPA guidelines, the use of the SANS is still viable for research and clinical practice. Furthermore, as proposed in the EPA guidelines, specific items which do not represent NS (like inappropriate affect and inattention) were not used in this study^32^. In addition, the SANS is a widely used instrument in clinical routine and a significant number of previous NS studies have used this scale allowing comparability of the present work with previous findings and facilitates transfer into clinical practice. Second, while this study addressed the main clinical factors contributing to secondary NS, it did not investigate other potential secondary sources such as environmental, psychological, and biological factors. Moreover, the dataset lacked detailed information regarding the number and frequency of psychotic episodes in the BD-I group. Understanding how the frequency of psychotic episodes might contribute to NS and neurocognitive deficits in BD-I patients could be a relevant avenue for future research. The cross-sectional study design with no follow-up visits examining changes of symptomatology over time does not allow a clear differentiation of primary and secondary NS. Future longitudinal studies will be necessary to further evaluate the role of primary and secondary NS on clinical outcome and function. Extrapyramidal side effects of antipsychotic medications are recognized as a cause of diminished expressivity, independent from efficacy and dosage of these medications^87^. Data on extrapyramidal symptoms were not available in our dataset, hindering their inclusion in the multivariate analysis. As a consequence, though antipsychotic medication dose was included in the analysis potential associations with extrapyramidal symptoms could not be fully excluded. The total score of the Hamilton Depression Scale does not fully disentangle depression from negative symptoms and could explain at least in part the observed overlap with anhedonia-asociality assessed with the SANS. Our cohort was not replicating age and sex related differences in negative symptoms and sex differences in working memory, as known from the existing literature. The association of age with negative symptoms^64^ remains controversial Some studies found no association with age or stability of negative symptoms over time^59,60,88^, others showed that NS were differently related to either older age^89^ or younger age^63^. In addition, longitudinal studies provided evidence of different NS trajectories with persistence in over 50%, thus suggesting that in some individuals NS might decline while in other they might persist or re-occur^59^. With respect to the age range, studies who directly addressed the relationship between age and NS included samples with larger age range^63,88,89^. In contrast, the relative narrower age range (35.9 ± 8.9; 21-50) in the present study might be a potential limitation to identify this association. There is consistent evidence that NS are known to be more severe in males^64–68^. Studying sex differences in NS was not an objective of the present study, and therefore the study cohort was not optimized for such an analysis. Therefore, the inequal sex distribution in the present cohort might the lack of relationship between sex and NS. Sex differences in working memory are reported in previous research, with men being more impaired than female^70–72^. However, the effect size of the difference was small in these studies, and large sample sizes and a design aimed to evaluate the sex influence on working memory were necessary for a positive result to emerge. In addition, no significant differences were found in other investigations^90–92^, or an inverse relationship in both SSD^73^ and BD-I^74^.

## CONCLUSIONS

Our findings confirm that NS are prevalent in outpatients with SSD and BD, highlighting the significant role of disorganization as a possible underlying source of avolition and anhedonia. The relationship between avolition-apathy and impaired WM suggests the presence of shared symptom-cognition relationships across SSD and BD-I. Collectively, these findings enhance our understanding of NS and their relationship to other clinical factors and cognitive impairments across the schizophrenia-bipolar spectrum. More broadly these findings might help to advance the development of targeted treatment and preventive measures for NS across SSD and BD.

## Data Availability

All data used in the presented study are available online at https://openfmri.org/dataset/ds000030/

## ACKNOWLEDGEMENTS

All data used in this study were derived from the UCLA CNP cohort. Data can be download from the publicly available database OpenfMRI (https://openfmri.org/dataset/ds000030/).

## CONFLICT OF INTERESTS

The authors declare that they have no conflicts of interest related to this study.

## Notes

### Competing Interest Statement

The authors have declared no competing interest.

### Funding Statement

This study did not receive any funding

### Author Declarations

The study used only openly available human data from the UCLA CNP that can be downloaded at: https://openfmri.org/dataset/ds000030/

### Summary of Updates

Introduction has been revised for clarity and to better explain the tested hypotheses. Methods have been revised to easier follow the methodological approach. Results have been revised and additional analyses have been performed to show robustness of the main findings.

