## Supplement for "Negative Symptoms and Their Associations with Other Clinical Variables and Working Memory Across the Schizophrenia Spectrum and Bipolar Disorder"

**SUPPLEMENTARY MATERIALS**

**Table S1**. DSM IV criteria of patients

| DSM IV code | Number of subjects |
| --- | --- |
| 295.10 Schizophrenia, Disorganized Type | 1 |
| 295.30 Schizophrenia, Paranoid Type | 21 |
| 295.60 Schizophrenia, Residual Type | 6 |
| 295.70 Schizoaffective Disorder | 11 |
| 295.90 Schizophrenia, Undifferentiated Type | 11 |
| 296.40 Bipolar I Disorder, Most Recent Episode Hypomanic | 4 |
| 296.41 Bipolar I Disorder, Most Recent Episode Manic, Mild | 2 |
| 296.42 Bipolar I Disorder, Most Recent Episode Manic, Moderate | 1 |
| 296.45 Bipolar I Disorder, Most Recent Episode Manic, In Partial Remission | 3 |
| 296.46 Bipolar I Disorder, Most Recent Episode Manic, In Full Remission | 6 |
| 296.51 Bipolar I Disorder, Most Recent Episode Depressed, Mild | 2 |
| 296.52 Bipolar I Disorder, Most Recent Episode Depressed, Moderate | 4 |
| 296.53 Bipolar I Disorder, Most Recent Episode Depressed, Severe Without Psychotic Features | 5 |
| 296.55 Bipolar I Disorder, Most Recent Episode Depressed, In Partial Remission | 8 |
| 296.56 Bipolar I Disorder, Most Recent Episode Depressed, In Full Remission | 5 |
| 296.62 Bipolar I Disorder, Most Recent Episode Mixed, Moderate | 1 |
| 296.64 Bipolar I Disorder, Most Recent Episode Mixed, Severe With Psychotic Features | 3 |
| 296.7 Bipolar I Disorder, Most Recent Episode Unspecified | 5 |

**Table S2**. mood stabilizer intake

|  | SSD | BD-I |
| --- | --- | --- |
| Lithium | 5 | 9 |
| Valproate | 5 | 9 |
| Lamotrigine | 2 | 10 |
| Topiramate | 1 | 6 |
| Oxcarbazepin | - | 5 |
| Carbamazepine | 1 | 1 |
| Gabapentine | 0 | 1 |

**Table S3**: relationship of age with negative symptoms and working memory

|  | Transdiagnostic | | | SSD | | BD-I | |
| --- | --- | --- | --- | --- | --- | --- | --- |
|  | *Rho** | *P* | *Rho** | | *P* | *Rho** | *P* |
| Avolition-apathy | 0.11 | 0.29 | 0.19 | | 0.19 | -0.05 | 0.72 |
| Anhedonia-asociality | 0.17 | 0.10 | 0.19 | | 0.19 | 0.22 | 0.13 |
| Alogia | -0.06 | 0.54 | -0.16 | | 0.27 | -0.06 | 0.68 |
| Blunted affects | 0.01 | 0.92 | 0.06 | | 0.69 | -0.16 | 0.27 |
| Working memory | -0.25 | 0.015 | -0.23 | | 0.10 | -0.19 | 0.20 |

*Spearman rank correlation test

**Table S4**: relationship of sex with negative symptoms and working memory

|  | Transdiagnostic | | SSD | | BD-I | |
| --- | --- | --- | --- | --- | --- | --- |
|  | *W** | *P* | *W** | *P* | *W** | *P* |
| Avolition-apathy | 1141 | 0.70 | 238 | 0.82 | 215 | 0.75 |
| Anhedonia-asociality | 1026 | 0.72 | 240 | 0.78 | 140 | 0.04 |
| Alogia | 1091 | 0.99 | 224 | 0.94 | 189 | 0.09 |
| Blunted affects | 1067 | 0.86 | 203 | 0.56 | 201 | 0.41 |
| Working memory | 948 | 0.29 | 239 | 0.82 | 224 | 0.9 |

*Wilcoxon rank-sum test. Statistically nominal significant results (uncorrected for multiple comparison) in red. No significant results after correction for multiple comparisons with the Benjamini-Hochberg procedure.

**Table S5.** Subgroup analyses of negative symptoms and potential sources of secondary negative symptoms in patients with SSD and BD-I

|  | Beta [95% CI] | | | | | |
| --- | --- | --- | --- | --- | --- | --- |
|  | *Avolition-Apathy* | *Anhedonia-Asociality* | *Alogia* | *Blunted affect* | *Amotivation* | *Diminished expression* |
| SCHIZOPHRENIA |  |  |  |  |  |  |
| Model fit | r^2^_adj_ = .24  F(4,45) =4.83** | r^2^_adj_ = .19  F(4,45) =3.81 | r^2^_adj_ = -.057  F(4,45) =0.34 | r^2^_adj_ = -0.006  F(4,45) =0.93 | r^2^_adj_ = .25  F(4,45) =5.19** | r^2^_adj_ = -0.04  F(4,45) =0.51 |
| Positive Symptoms | -0.04[-0.20 – 0.12] | -0.014[-0.18 – 0.15] | 5.9^-3^[-0.14 – 0.15] | 0.03[-0.10 – 0.16] | -0.05[-0.35 – 0.24] | 0.04[-0.24 – 0.32] |
| Disorganization | 0.32[0.14 – 0.50]*** | 0.2[0.02 – 0.39]* | -0.04[-0.12 – 0.21] | -0.03[-0.11 – 0.14] | 0.53[0.20 – 0.86]** | 0.01[-0.30 – 0.33] |
| Depression | 0.04[-0.007 – 0.09] | 0.05[0.004 – 0.10]* | <0.001[-0.043 – 0.043] | 0.03[-0.008 – 0.05] | 0.09[0.006 – 0.18]* | 0.03[-0.05 – 0.12] |
| RSP-eq dose | 0.02[-0.02 – 0.07] | 0.03[-0.01 – 0.08] | 2.00^-2^[-0.02 – 0.06] | 0.01[0.005 – 0.05] | 0.06[-0.02 – 0.14] | 0.03[-0.05 – 0.10] |
| BIPOLAR DISORDER |  |  |  |  |  |  |
| Model fit | r^2^_adj_ = .0055  F(4,42) =1.06 | r^2^_adj_ = .19  F(4,41) = 3.58* | r^2^_adj_ = -.04  F(4,42) =0.55 | r^2^_adj_ = .13  F(4,42) =2.75* | r^2^_adj_ = .06  F(4,41) =1.73 | r^2^_adj_ = .04  F(4,42) =1.5 |
| Positive Symptoms | 0.38[-0.15 – 0.92] | -0.16[-0.53 – 0.21] | -0.05[-0.17 – 0.06] | 0.10[-0.19 – 0.39] | 0.27[-0.53 – 1.07] | 0.04[-0.32 – 0.42] |
| Disorganization | 0.13[-0.18 – 0.43] | 0.18[-0.03 – 0.40] | -0.02[-0.09 – 0.04] | -0.001[-0.16 – 0.16] | 0.34[-0.11 – 0.80] | -0.02[-0.23 – 0.19] |
| Depression | -0.02[-0.07 – 0.04] | 0.05[0.01 – 0.09]* | 0.005[-0.008 – 0.018] | 0.006[-0.025 – 0.04] | 0.03[-0.06 – 0.11] | 0.01[-0.029 – 0.051] |
| RSP-eq dose | 0.01[-0.03 – 0.05] | 0.004[-0.02 – 0.03] | -0.001[-0.01 – 0.007] | 0.035[0.01 – 0.06]** | 0.01[-0.05 – 0.08] | 0.04[0.005 – 0.06]* |

Group specific linear regressions of negative symptoms and potential sources of secondary negative symptoms. Linear multivariate regressions of 4 negative symptom domains as dependent variables with potential sources of secondary negative symptoms as independent variables for SSD and BD-1 patients independently. Beta values and corresponding 95% confidence intervals are given; Positive symptoms were measured using the Scale for the assessment of positive symptoms (SAPS); Depressive symptoms were measured using the Hamilton Depression Scale (21 items); RSP-eq dose, daily antipsychotic dose as risperidone equivalent; *p<0.05 **p<0.005 ***p<0.001

**Table S6**. Relationship between negative symptoms domains and other clinical factors, results corrected for age and sex

|  | Beta [95% CI] | | | |
| --- | --- | --- | --- | --- |
|  | Avolition-Apathy | Anhedonia-Asociality | Alogia | Blunted affect |
| Model fit | r^2^_adj_ = .18  F(7,89) = 3.92*** | r^2^_adj_ =.25  F(7,88) =5.48*** | r^2^_adj_ = .14  F(7,89) =3.24** | r^2^_adj_ = .13  F(5,91) =3.80* |
| Group | -0.55 [-1.39 – 0.28] | -0.59 [-1.34 – 0.15] | -0.74[-1.29- -0.19]** | -0.53 [-1.24 – 0.17] |
| Age | 0.005 [-0.03-0.04] | 0.01 [-0.02 – 0.04] | -0.011 [-0.03-0.01] | -0.01 [-0.04-0.01] |
| Sex | 0.11[-0.50-0.77] | 0.09 [-0.47 – 0.66] | 0.13[-0.29 – 0.54] | 0.25 [-0.28-0.79] |
| Positive Symptoms | 0.007 [-0.14 – 0.16] | -0.03 [-0.16 – 0.10] | 0.009[-0.09 – 0.11] | 0.03 [-0.09 – 0.17] |
| Disorganization | 0.25[0.11 – 0.40]*** | 0.20 [0.07 – 0.34]** | 0.04[-0.05 – 0.14] | 0.02 [-0.10 – 0.15] |
| Depression | 0.02 [-0.012 – 0.05] | 0.05 [0.02 – 0.08]*** | 0.0001[-0.02 – 0.02] | 0.02[-0.007 – 0.05] |
| RSP-eq dose | 0.02 [-0.01 – 0.05] | 0.01 [-0.01 – 0.04] | 0.009[-0.01 – 0.03] | 0.03[0.006 – 0.06]* |

Linear multivariate regressions of 4 negative symptom domains as dependent variables with potential sources of negative symptoms as independent variables across SSD and BD-I patients. Beta values and corresponding 95% confidence intervals are given; Positive symptoms were measured using the Scale for the assessment of positive symptoms (SAPS); Depressive symptoms were measured using the Hamilton Depression Scale (21 items); RSP-eq dose, daily antipsychotic dose as risperidone equivalent; *p<0.05 **p<0.005 ***p<0.001. Statistically significant results after correction for multiple testing (Beniamini-Hochberg procedure, false discovery rate 5%) are bolded.

**Table S7**. Relationship between clinical variables and working memory, results corrected for sex

|  | Transdiagnostic | Schizophrenia | Bipolar disorder |
| --- | --- | --- | --- |
| Model fit | r^2^_adj_ = .26  F(10,85) = 4.39*** | r^2^_adj_ = -.05  F(9,40) = 0.74 | r^2^_adj_ = .27  F(9,36) = 2.86* |
| Group | 2.76[1.11 – 4.41]** | / | / |
| Sex | 0.06 [-1.15-1.26] | -0.18 [-2.29 – 1.93] | -0.77 |
| Avolition-Apathy | -0.74[-1.22 - -0.26]** | -0.57[-1.39 – 0.26] | -1.08[-1.64 – -0.51]*** |
| Anhedonia-Asociality | 0.46[-0.10 – 1.03] | 0.002[-0.85 – 0.86] | 1.37[0.44 – 2.29]** |
| Alogia | -0.40[-1.19 – 0.39] | -0.46[-1.48 – 0.56] | 1.37[-1.67 – 4.40] |
| Blunted affect | -0.05[-0.65 – 0.56] | 0.03[-0.88 – 0.94] | -0.51[-1.64 – 0.62] |
| Positive symptoms | 0.17[-0.11 – 0.46] | 0.11[-0.24 – 0.47] | 0.56[-0.31 – 1.43] |
| Disorganization | -0.015 [-0.32 – 0.29] | 0.10[-0.36 – 0.56] | -0.13[-0.59 – 0.34] |
| Depression | -0.008[-0.08 – 0.06] | 0.04[-0.08 – 0.16] | -0.09[-0.19 – 0.02] |
| RSP-eq dose | -0.022[-0.08 – 0.03] | 0.03[-0.07 – 0.127] | -0.05[-0.12 – 0.02] |

*Linear regressions of negative symptom domains and other clinical variables as independent variables and working memory sum score as dependent variable across SSD and BD-1 patients. Beta values and corresponding 95% confidence intervals are given; Positive symptoms were measured using Scale for the assessment of positive symptoms (SAPS); depressive symptoms were measured using Hamilton Depression Scale (21 items); RSP-eq dose, daily antipsychotic dose as risperidone equivalent; *p<0.05 **p<0.005 ***p<0.001. Statistically significant results after correction for multiple testing (Beniamini-Hochberg procedure, false discovery rate 5%) are bolded.*
